# Modelling the epidemiological trend and behavior of COVID-19 in Italy

**DOI:** 10.1101/2020.03.19.20038968

**Authors:** Alessandro Rovetta, Akshaya Srikanth Bhagavathula, Lucia Castaldo

**Author notes:** **Corresponding author** Alessandro Rovetta, Mensana srls research and disclosure division, Via Moro Aldo 5 - 25124 Brescia, Italy., ORCID: http://orcid.org/0000-0002-4634-279X.

## Abstract

As of May 14, 2020, Italy has been one of the red hotspots for the COVID-19 pandemic. With over 220,000 confirmed cases and almost 33,000 confirmed deaths reported from February, it is necessary to fully understand the spread of COVID-19 in this country. By S.E.I.R. simulation, we estimated the most representative basic reproduction number R0 for the three most affected regions from February 22 to March 14, 2020. In doing so, we have been able to evaluate the consistency of the first containment measures until the end of April, as well as identify possible SARS-CoV-2 local behavior mutations and specificities. Next to that, through new estimates of the infection mortality rate, we recalculated a more plausible number of real infected. Finally, given the absolutely anomalous trend of the Lombardy region, we looked for correlations between COVID-19 total cases and air pollutants such as PM 10 and PM 2.5.

## Introduction

The current surge of COVID-19 pandemic is devastating globally, with over 4,200,000 cases and more than 290,000 deaths reported [3]. In Europe, COVID-19 cases have started to dramatically increase from the first week of March 2020. Of these, Italy was grappling with the worst outbreak, with over 35,713 confirmed cases and around 3000 confirmed deaths by March 18, 2020 [1]. This exponential increase in COVID-19 positive cases in Italy raised turmoil, and the government decree to a lockdown of the entire country [2].

In this research, making use of the epidemic parameters provided by WHO, we utilized the S.E.I.R. mathematical model to predict the trend of infections during the first half of March 2020, when the effects of the lockdown were not yet measurable. This allowed us to signal the presence of SARS-CoV-2 local behavior changes due to possible evolutionary genetic mutations, correlations with pollution like PM 10 and PM 2.5, mismanagement of the crisis by national government agencies, non-compliance with the lockdown rules by citizens or other unknown factors. To do this, it was sufficient to compare the general trend foreseen by the S.E.I.R. with the estimated one as well as highlight the discrepancies between the individual Italian regions.

## Methods

To carry out this study, the most recent data found in the scientific literature relating to COVID-19 total and active cases, deaths, recoveries, and all epidemic parameters, have been used [1]. We focused especially on Lombardy since it was by far the most afflicted region in Italy. Considering the novel coronavirus incubation period is around 3 – 6 days, with a range from a minimum of 2 days to a maximum of 14, we firstly examined the population of COVID-19 cases reported between February 22^th^ and March 14^th^, 2020 [4]. Following this, we analyzed the Pearson linear correlation between the number of COVID-19 total cases and the concentrations of PM 10 and PM 2.5. All daily PM data were collected from the ARPA regional websites in the interval January 1^st^ - May 14^th^. Several monitoring stations were used and all results were provided with a Gaussian 95% confidence interval. All data have been organized into two time periods such as the “pre-lockdown”, from January 1st until February 29th, and the “post-lockdown”, from March 1st until May 14^th^; this served us to estimate the link between the virus spread and the particulate matter (as emissions dropped dramatically during the lockdown). Other types of correlation with population density and number of inhabitants were also investigated.

### S.E.I.R. Modelling

Assuming true the probable “non-relapse patients” hypothesis, we applied the S.E.I.R. model to predict the novel coronavirus evolution in Italy as it is suitable for describing the spread of a virus in populations where no restrictions have been applied [5]. Thanks to the comparison between the S.E.I.R. values and the theoretical estimates (TE) in the short period, it is likely to highlight essential behavior mutations and/or containment strategies effectiveness. We used S.E.I.R. differential equations and non-linear methods to resolve the gaps analytically [6]. We examined Lombardy, Emilia Romagna, and Piedmont separately because of the huge discrepancy of COVID-19 cases between these and other Italian regions. An iterative algorithm was developed using C++ programming language to find a solution through a finite discretization method [Appendix 2]. Given the very low deaths-population ratio, the total population number has been considered constant.

### Software iterative algorithm

By entering the initial values for the incubation time 1/σ, the recovery time 1/γ, the basic reproduction number R0, the number of infected *I*_0_, and the number of recovered *R*_0_ on February 22, 2020, the software prints the S.E.I.R. predictions day by day. The best epidemic parameters were estimated through continuous iteration until the “closest values to the real ones” were reached until March 12, 2020. The number of initial incubates was calculated with the formula E_0_ = R0·I_0_. We report below the system of equations and their discretization through the finite increment δ*t*:

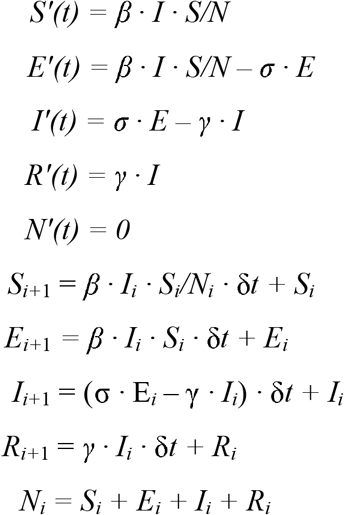

· 1/γ is the incubation time;

· 1/σ is the recovery time;

· R0 is the basic reproduction number;

· *S* is the number of susceptible people;

· *E* is the number of active exposed people (people in incubation);

· *I* is the number of active infected people;

· *R* is the number of recovered people (no longer infectable).

### R0 statistical analysis

The compatibility between S.E.I.R. predictions *x_i_* and TE values *X_i_* was investigated within a closed ball of radius 5 days centered in March 7 (from March 2 to March 12, 2020). We searched for the best infection mortality rate *m* and basic reproduction number R0 through the minimization of two estimators: the first, called Δ, was the algebraic mean value of the absolute percentage differences *δ_i_* between the best value *X_i_* for the *x_i_* S.E.I.R. value Gaussian distribution *G* (*X_i_*, *σ_i_)* and the *x_i_* value itself, according to the formula *δ_i_* = |*X_i_* - *x_i_* | / *x_i_*. This allows us to assess the quality of the S.E.I.R. modeling. The second, called *ε*, was the algebraic mean value of the ratios *β_i_* between the fixed standard deviation 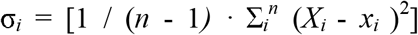 and the corresponding *x_i_*, according to the formula *β_i_* = *σ_i_* / *x_i_*. This allow us to calculate the *x_i_* relative errors. The lower the Δ, the more representative R0 is of the TE evolution; the lower the *ε*, the more accurate is the TE. The iteration was carried out until *ε* and Δ minimums were reached. The finite increments chosen for *m* and R0 were *δm* = 0.2 and *δ*R0 = 0.1 respectively. Every combination with *m* in [0.7, 1.5] and R0 in [1, 7] was tried. The chosen significance limit for *ε* and Δ was 0.1 (i.e. Δ and *ε* must be lower than or equal 0.1 for a result to be acceptable); the iteration ended when Δ and *ε* < 0.05. The best R0 confidence intervals were calculated considering the Gaussian distribution *G* (R0, 2·*δ*R0). We reported a range interval “CRI” for all compatibles R0 we found. All the analysis was carried out through our C ++ software and Microsoft Excel. Since *m* is subject to a very wide margin of error, when more than one (m, R0) compatible couple was computed, we utilized the weighted average 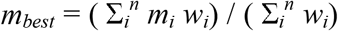, with 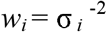.

### COVID-19 real cases estimate

In order to calculate the real number of COVID-19 total cases in Lombardy, we used an estimation method we called “Theoretical Estimate” (TE). Thanks to the results of another study, in which the number of regional and national deaths until May 5, 2020, was compared with those of the previous 5 years, it was possible to calculate the number of theoretical cases by adding the number of COVID-19 missing cases until the expected infection mortality was achieved [7]. To do this, we considered a mortality rate free to vary in the range [0.6, 1.6] [7 – 9]. The “COVID-19 total deaths number” was shifted 7 days backward due to the time between contracting the disease and demise. We also calculated the difference between confirmed and estimated COVID-19 trough the ratio *α* = estimated cases / confirmed cases.

### PM 10 data analysis

First, we collected PM 10 daily averages data on the most affected cities of the three main regions involved in the COVID-19 epidemic, such as Lombardy, Piedmont and Emilia Romagna; then, we did the same for some regions where the novel coronavirus spree was not so pressing, like Lazio and Campania. Finally, we built the symmetric correlation matrix *ρ_ij_*

|  |  |  |  |
| --- | --- | --- | --- |
| 1 |  |  |  |
| $\rho_{12}$ | 1 | | |
| $\rho_{13}$ | $\rho_{23}$ | 1 | |
| $\rho_{14}$ | $\rho_{24}$ | $\rho_{34}$ | 1 |

where

i. ρ_12_ is the PM 10 – COVID-19 confirmed cases Pearson correlation index
ii. ρ_13_ is the PM 10 – population density Pearson correlation index
iii. ρ_14_ is the PM 10 – total population Pearson correlation index
iv. ρ_23_ is the COVID-19 cases – population density Pearson correlation index
v. ρ_24_ is the PM 10 – COVID-19 Pearson correlation index
vi. ρ_34_ is the population density – total population Pearson correlation index

However, since the first outbreak occurred in northern Italy, it is plausible to think the infection did not have the same contagion power in the southern regions; thus, we recalculated the matrix *ρ* only for the three most infected regions to check whether there was a local correlation. Finally, in order to make the investigation even more precise and specific, we repeated the operation once again to all Lombardy provinces. All the values *ρ_ij_* have been reported with their relative p-values, according to the form *ρ_ij_* (p-value*_ij_*). All the PM 10 average daily values were reported with Gaussian 95% confidence intervals (*AV*-2·σ/√*N*, AV + 2·σ/√*N)*.

### PM 2.5 data analysis

Since other studies have been conducted on the PM 2.5 – novel coronavirus correlation at national level, we have focused exclusively on the Lombardy region, analyzing the data of all the monitoring units of all the provinces through the previous defined ρ correlation matrix [10].

## Results

### Epidemic forecast

For each infection mortality rate *m* a compatible value of R0 was found; therefore, we utilized the weighted average *m* = 0.011 (95% CI: 0.006 - 0.016). For the Lombardy region in the period February 22 - March 12, 2020, using the epidemic parameters 1/*γ* and 1/*σ* provided by WHO, we estimated a basic reproduction number R0 = 3.91 (95% CI: 3.87 - 3.94, CRI: 3.82 - 3.91), with Δ = 0.02 (95% CI: 0.01 - 0.03) and *ε* = 0.04 (95% CI: 0.03 - 0.05). The estimated number of real infections exceeds that of confirmed infections by a factor *α* ~ 34 until May 1, 2020 (*α* = 34.1, 95% CI: 33.0 - 35.3). The separation point between S.E.I.R. and TE trends is positioned in a neighborhood of March 12, 2020; ergo, that is the period when the lockdown began to take effect [Figure 1]. The two corner points I and II in figure 1 indicate further decreases in R0. About Emilia Romagna, we estimated a basic reproduction number R0 = 2.22 (95% CI: 2.18 - 2.26, CRI: 2.20 - 2.23), with Δ = 0.09 (95% CI: 0.07 - 0.13) and *ε =* 0.10 (95% CI: 0.08 - 0.12) in the same Lombardy investigation period. At May 1, 2020, the estimated number of Emilia Romagna real infections exceeds that of confirmed infections by a factor *α* ~ 24 (α = 23.7, 95% CI: 23.3 - 23.9). The substantial differences between the Lombardy and Emilia Romagna R0s show the behavior of SARS-CoV-2 is local i.e. it is potentially linked to demography, air quality, genetic mutations and other factors. Furthermore, in Emilia Romagna the novel coronavirus seemed to have circulated longer naturally (as if the lockdown was not in progress) however causing less serious damage than in Lombardy. In particular, the corner point I in figure 2 signals an R0 increase which lasted approximately until March 21^st^ [Figure 2]. As for Piedmont, we estimated a R0 = 2.52 (95% CI: 2.48 - 2.56), with Δ = 0.07 (95% CI: 0.01 - 0.13) and *ε* = 0.07 (95% CI: 0.06 - 0.09) from February 29 to March 14, 2020. In fact, Ministry of Health data show the infection appears to have started about a week late in this region. Moreover, an important slope increase has occurred at March 14 (15^th^ day) [Figure 3]; this remained almost constant until April 4 (corner point II). On April 25, the estimated COVID-19 total cases in Piedmont exceed those confirmed by a factor *α* = 23 (α = 22.5, 95% CI: 22.0 - 23.0). Between March 23 - 24, we begin to notice the first real positive effects of the lockdown.

**Figure 1.**
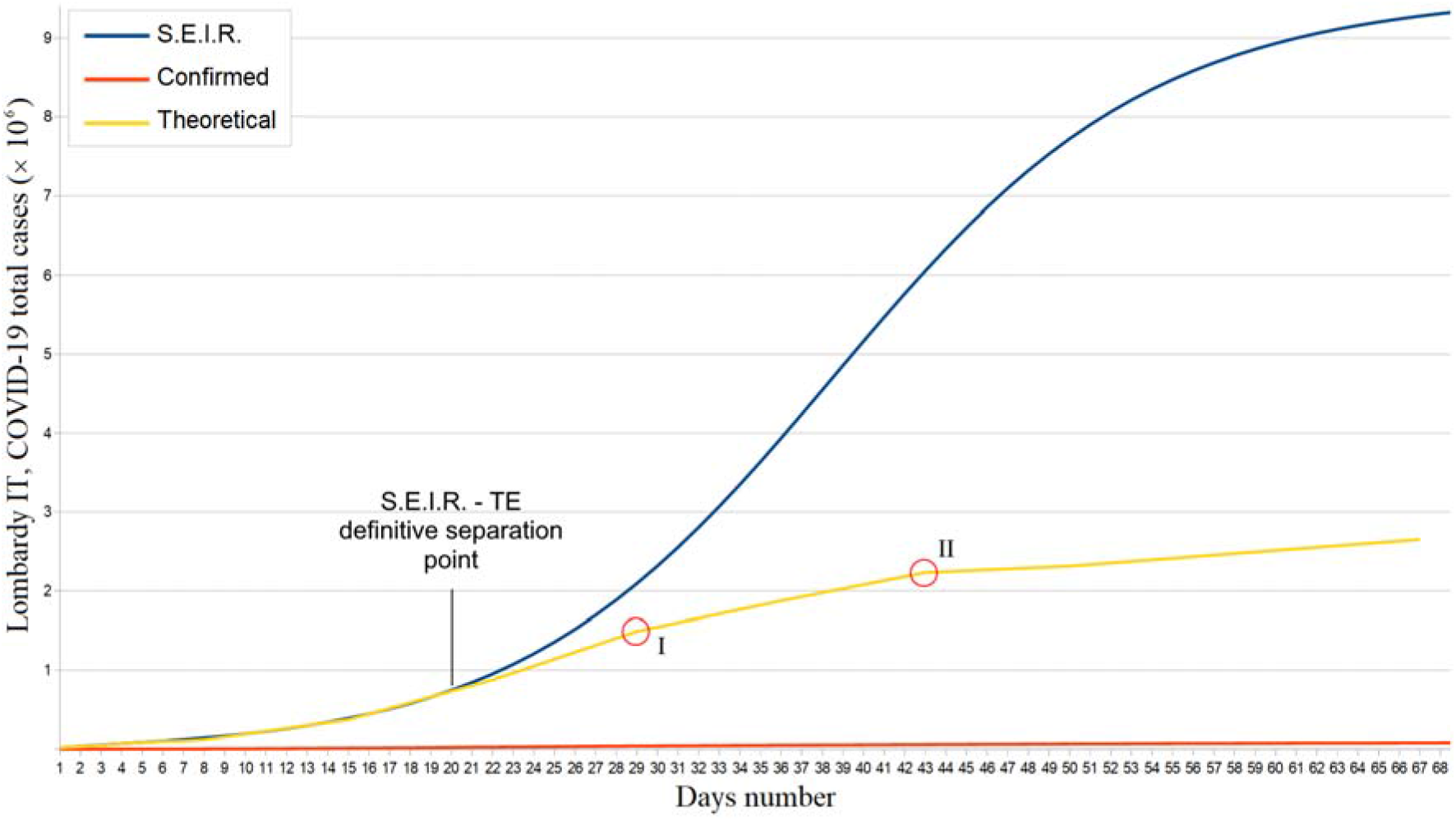
Lombardy confirmed, theoretical, and S.E.I.R. simulation total COVID-19 cases trends from February 22 to May 1, 2020.

**Figure 2.**
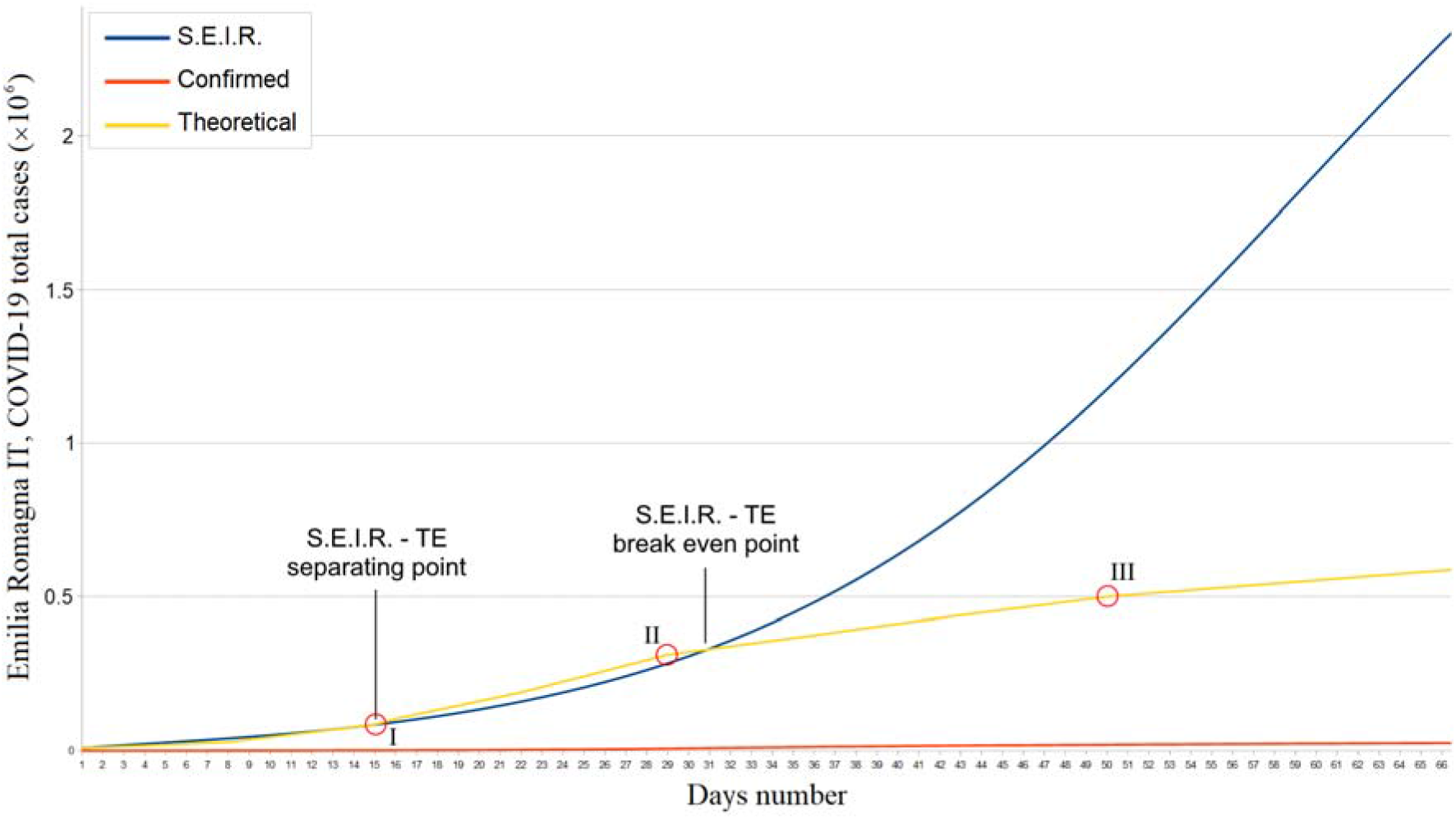
Emilia Romagna confirmed, theoretical, and S.E.I.R. simulation total COVID-19 cases trends from February 29 to April 30, 2020.

**Figure 3.**
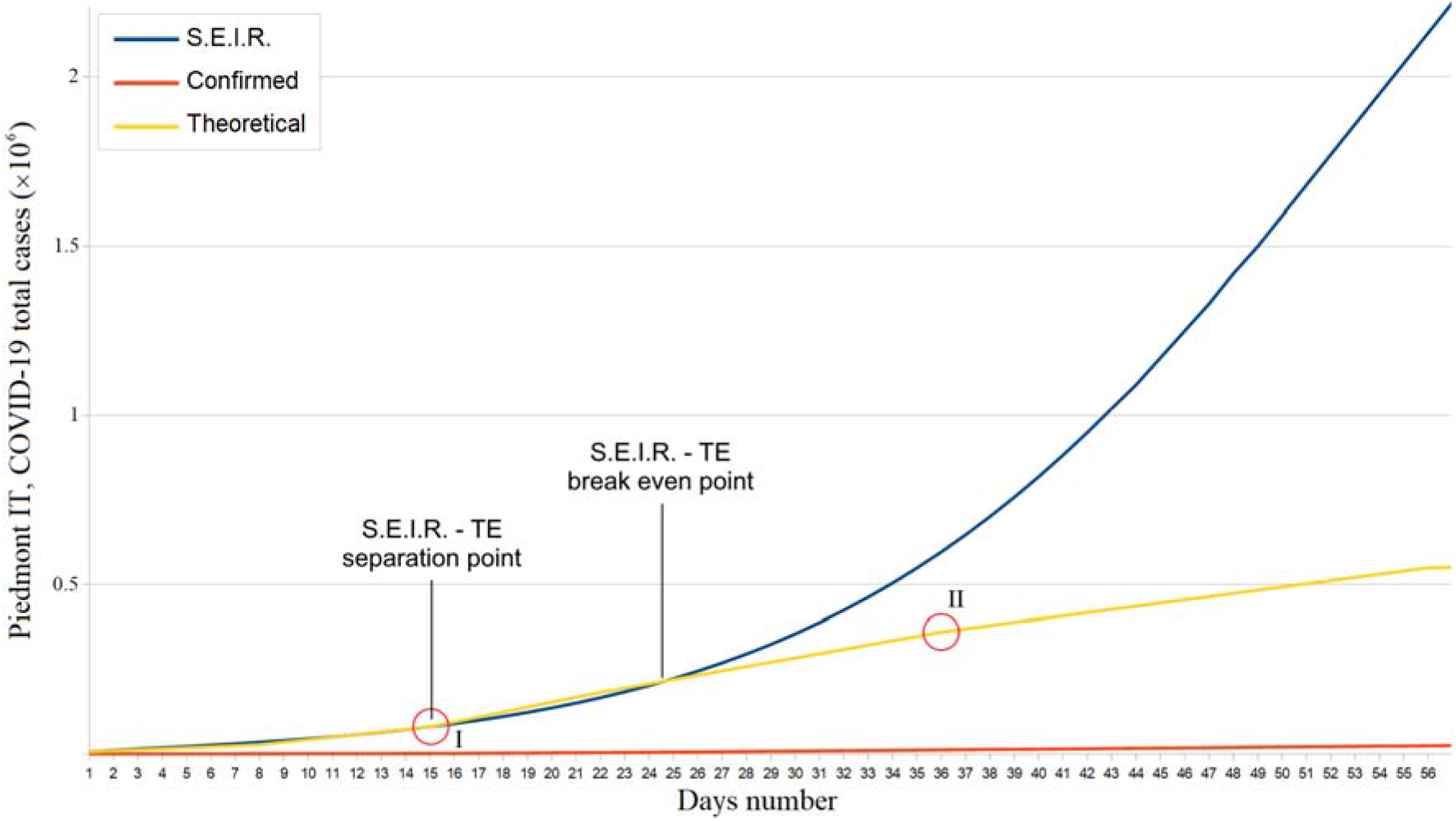
Piedmont confirmed, theoretical, and S.E.I.R. simulation total COVID-19 cases trends from February 29 to April 25, 2020.

### Pollution correlation

No significant correlation between SARS-CoV-2 and PM 10 was found. In particular, neither at national nor local level, a p-value < .1 has been reached and the higher Pearson index achieved was ρ = .34 [Tables 1 and 2]. Despite this is not enough to completely rule out any link between the novel coronavirus and this type of pollution, and ulterior investigations must be carried out at the molecular level, this allows us to state that PM 10 certainly cannot be the only discriminant characterizing the SARS-CoV-2 spread and virulence. On the contrary, a moderate correlation between SARS-CoV-2 spread and PM 2.5 was highlighted in Lombardy during the first two weeks of March, with a p-value = .07 and a Pearson correlation coefficient ρ = .56 [Tables 3 and 4]. Beyond that time, the above correlation has decreased in favor of a stronger SARS-CoV-2 – provinces population number correlation (p-value < .0001, ρ = 0.9) [Figure 4]. This indicates a very large fraction of the total Lombardy population has been infected by the virus. At the same time, no correlation between PM 2.5 and the other measured quantities was found. As confirmation of the lower virus contagiousness and spread in Emilia Romagna, the SARS-CoV-2 – provinces population number correlation grew much more slowly than in Lombardy, reaching its maximum value on 14 May, 2020 (ρ = .57, ρ-value = .11). Finally, in Piedmont there was a strong correlation between the COVID-19 cases and the population density (ρ = 0.71, p-value = 0. 048 on May 14, 2020) unlike Emilia Romagna and Lombardy [Appendix 1]. No SARS-CoV-2 – province population number was found (p-value >> 0.1).

**Table 1.** Top Italian “PM 10 polluted and COVID-19” affected cities.

| Region | City | pre-lockdown (Jan 1, Feb 29) |  | lockdown (March 1, May 14) |  | Population Density (P/km<sup>2</sup>) | Population (x 10<sup>6</sup>) |
| --- | --- | --- | --- | --- | --- | --- | --- |
|  |  | PM10 AV (µg/m<sup>3</sup>) | 95% CI (SEM) | PM10 AV (µg/m<sup>3</sup>) | 95% CI (SEM) |  |  |
| Lombardy | Milan | 56 | 50 - 62 | 26 | 23 - 29 | 2063 | 3.209 |
|  | Brescia | 50 | 46 - 55 | 27 | 24 - 30 | 264.5 | 1.264 |
|  | Bergamo | 43 | 38 - 47 | 21 | 19 - 24 | 417.9 | 1.108 |
| Emilia Romagna | Reggio Emilia | 54 | 49 - 59 | 26 | 22 - 29 | 232.2 | 0.533 |
|  | Piacenza | 52 | 47 - 57 | 24 | 21 - 27 | 111.1 | 0.287 |
|  | Bologna | 46 | 40 - 51 | 21 | 18 - 24 | 2773 | 1.006 |
| Piedmont | Turin | 59 | 51 - 67 | 20 | 18 - 22 | 331 | 2.282 |
|  | Alessandria | 60 | 54 - 66 | 26 | 23 - 30 | 118.4 | 0.429 |
| Lazio | Roma | 40 | 38 - 41 | 23 | 22 - 24 | 810 | 4.354 |
|  | Roma Tiburtina | 51 | 48 - 54 | 21 | 19 - 22 | 3600 | 0.17 |
|  | Frosinone | 51 | 48 - 55 | 22 | 21 - 23 | 153.3 | 0.496 |
| Campania | Napoli | 50 | 48 - 51 | 27 | 26 - 27 | 2617 | 3.117 |

**Table 2.**
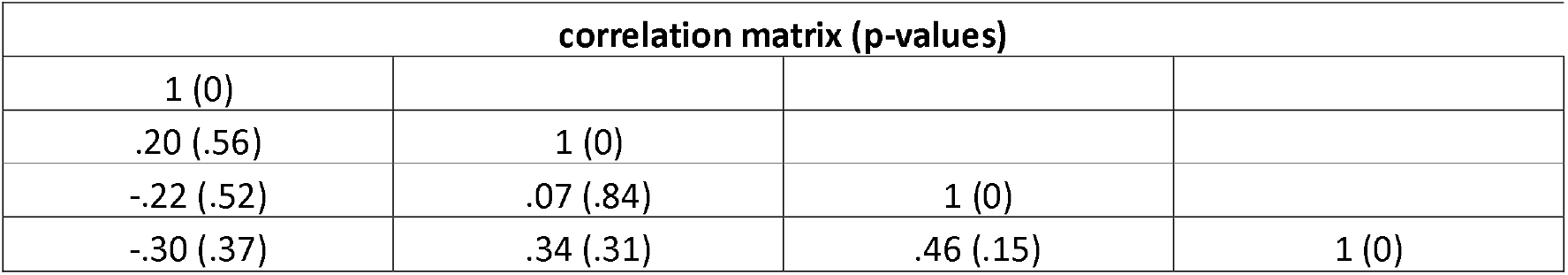
May 14. 2020. Top Italian “PM 10 polluted and COVID-19” affected cities correlation matrix.

| correlation matrix (p-values) |  |  |  |
| --- | --- | --- | --- |
| 1 (0) |  |  |  |
| .20 (.56) | 1 (0) |  |  |
| -.22 (.52) | .07 (.84) | 1 (0) |  |
| -.30 (.37) | .34 (.31) | .46 (.15) | 1 (0) |

**Table 3.** Lombardy cities populations, PM 2.5 daily average values from January 1 to May 14. 2020, and COVID-19 total cases until May 14, 2020.

| Cities | PM 2.5<br>($\mu\text{g}/\text{m}^3$) | 95% CI<br>(SEM) | COVID-19<br>cases (May 14) | Population<br>Density (P/$\text{km}^2$) | Total Population<br>($\times 10^6$) |
| --- | --- | --- | --- | --- | --- |
| Milan | 43 | 39 - 46 | 22151 | 2063 | 3.209 |
| Brescia | 41 | 39 - 44 | 14147 | 292.6 | 1.264 |
| Bergamo | 42 | 39 - 45 | 12443 | 264.5 | 1.108 |
| Cremona | 47 | 44 - 51 | 6323 | 204.5 | 0.362 |
| Monza B. | 44 | 41 - 47 | 5287 | 417.9 | 0.872 |
| Pavia | 45 | 41 - 49 | 4979 | 742 | 0.89 |
| Como | 39 | 36 - 42 | 3629 | 418.4 | 0.339 |
| Varese | 32 | 30 - 35 | 3379 | 2228 | 0.6 |
| Lodi | 39 | 37 - 41 | 3351 | 292.6 | 0.182 |
| Mantova | 39 | 37 - 41 | 3291 | 3729 | 0.872 |
| Lecco | 24 | 21 - 26 | 2645 | 177.3 | 0.415 |
| Sondrio | 21 | 19 - 22 | 1367 | 184.7 | 0.548 |

**Table 4.** Lombardy cities populations, PM 2.5 daily average values, and COVID-19 total cases: most significant correlation values and days.

| Date | correlation matrix (p-values) |  |  |  |
| --- | --- | --- | --- | --- |
| March 4. 2020 | 1 (0) |  |  |  |
|  | **.56 (.07)** | 1 (0) |  |  |
|  | -.08 (.82) | -.33 (.32) | 1 (0) |  |
|  | -.32 (.34) | .11 (.75) | .32 (.34) | 1 (0) |
| May 14. 2020 | 1 (0) |  |  |  |
|  | .46 (.13) | 1 (0) |  |  |
|  | .07 (.83) | .09 (.78) | 1 (0) |  |
|  | .30 (.34) | **.90 (.0001<)** | .35 (.26) | 1 (0) |

**Figure 4.**
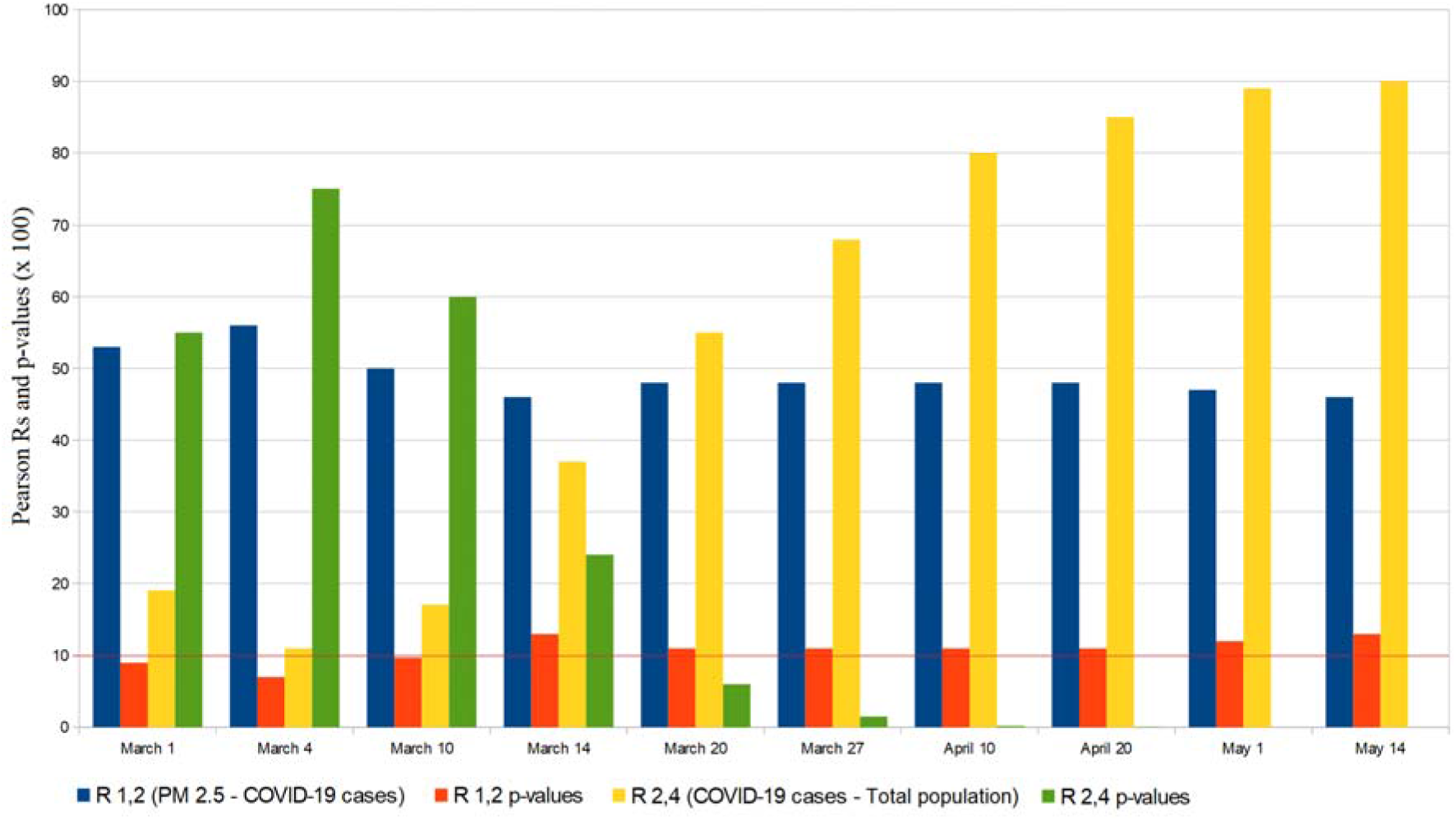
Lombardy cities “PM 2.5 aerosols and populations” correlations with COVID-19 cases.

### Further results

The matrices shown in table 4 are greatly in agreement with our COVID-19 total and active cases estimates: in fact, the correlation between PM 2.5 and the novel coronavirus spread in Lombardy was significantly moderate in the first two weeks of March; then, it decreased in correspondence with the rising difference between the S.E.I.R. predictions and the TEs. Moreover, the clear increase in the correlation between the number of COVID-19 total cases and the number of inhabitants of the various provinces of Lombardy supports and motivates what is shown in figure 1. However, the moderate correlation with PM 2.5 cannot be the only cause for the significant discrepancy of COVID-19 total cases between Lombardy and the other Italian regions: in fact, Emilia Romagna has an average PM2.5 concentration of 30 μg / m^3^ against the 35 μg / m^3^ of Lombardy, while Piedmont even reaches 39 μg / m^3^ [10]. In addition, no significant COVID-19 rising cases – PM 2.5 correlation was found in Piedmont: in fact, the COVID-19 total cases curve reached a steep slope much later than the other two regions and in a period in which the quantity of PM air pollutants was very low. For example, Turin outbreak became important around March 15, despite the PM 2.5 concentrations in the previous 15 days were < 20 μg / m^3^ [17]. Finally, the age groups of Lombardy and Emilia Romagna are demographically comparable i.e. the greater virulence of SARS-CoV-2 in the first cannot be explained through this data [27, 28]. For all these reasons, it is reasonable to assume the following scenarios:

I. SARS-CoV-2 has undergone evolutionary and anti-evolutionary genetic mutations [18 – 25]. Anyway, there are some research in contrast with this hypothesis [22].
II. lockdown management has been much more effective in some regions than in others;
III. the location of the outbreaks played a large role in the spread of SARS-CoV-2;
IV. Other factors speed up the spread of SARS-CoV-2 [26].

## Discussion

For this analysis, some educated guesses were made:

- given the antibody response identified in COVID-19 patients it seems unlikely the novel coronavirus, without significantly changing, could infect a patient again (in the short term) [11, 12];
- the mortality rate is too low to affect the evolution of the S.E.I.R. system [7–9];
- Young people and children appear to have an important role in the spread of infection [13–15],

Therefore, to analyze the SARS-CoV-2 dynamics in the initial stages, we have adopted the S.E.I.R. model (Susceptible → Exposed → Infected → Recovered) since it is suitable for describing the spread of a virus in a non-relapse free population. This allowed us to evaluate the effectiveness of the containment measures and/or any virus behavior mutation by comparing S.E.I.R. and theoretical trends [Figures 1 and 2]. Given the absolutely abnormal number of COVID-19 infections and deaths, we treated Lombardy as a stand-alone case; moreover, the modeling we have made concerns exclusively northern Italy since it was the most COVID-19 affected region. Our results show the effectiveness of the Italian lockdown. However, the strong correlation between the total infected and the number of inhabitants in Lombardy suggests the virus nevertheless circulated in a very substantial way among this region i.e. containment measures are likely to have been taken with a heavy delay. In fact, as of May 1, the estimated COVID-19 total cases in Lombardy were almost 3 million. Furthermore, it is plausible the effect of such a latency was aggravated by the presence of PM 2.5 as shown in figure 4. In this regard, we must point out the COVID-19 cases – PM 2.5 correlation p-value has never exceeded a maximum of .16 and the violation of the significance threshold does not always imply a lack of correlation (and vice versa) [16]. The substantial difference between the basic reproduction number of Lombardy and that of the other two most affected regions is the main symptom of a local behavior of the novel coronavirus. About this aspect, various speculations, hypotheses and theories were made: some of them concern a possible evolutionary genetic mutation of SARS-CoV-2 in Italy, which has made it more contagious or virulent [18 – 20]; if so, according to what has been pointed out, this should have afflicted Lombardy. Other researches instead assert the virus genetic mutations did not cause mutation in its behavior [22]; if so, the greater Lombard virulence should find explanations in correlation with pollution, delay in the lockdown or other factors, such as a major predisposition of the subjects to be infected and develop sever symptoms [10, 23]. However, it must be considered that many of these scenarios could prove to be true simultaneously. One hypothesis to be excluded is the bijective relation between local demography and SARS-CoV-2 spread: in fact, Lombardy and Emilia Romagna age groups are totally comparable [27, 28]. The behavior of the novel coronavirus in Piedmont was also peculiar: first of all, despite what happened in Lombardy and Emilia Romagna, we found a strong correlation between the population density and the number of total cases. Furthermore, the epidemic seems to have started with a delay of one week. This fact cannot be explained by the presence of particulate pollution since the concentrations of PM 2.5 and PM 10 were already below the safety threshold due to the quarantine. Therefore, it is possible this episode is linked to virus mutations, people lockdown violations, or other unknown factors. Other dynamics relating to work and university travel must be investigated. As for a possible link between PM 10 and novel coronavirus, we found no significant correlation. At national level, concentrations of PM 10 in some cities of central and southern Italy were comparable to those of Lombardy and Emilia Romagna. In particular, cities like Frosinone, Rome Tiburtina and Naples, where COVID-19 infections were few, had higher values than cities like Brescia and Bergamo where the infection was devastating. Furthermore, PM 10 concentrations in Emilia Romagna and Piedmont are equivalent to Lombard ones unlike the density of COVID-19 cases. Evaluating the hypothesis the first outbreak location was highly incident on the virus spread, we conducted the analysis in the Lombardy region alone but the result was still negative. The lack of a clear correlation, however, is not sufficient to exclude any relation between PM 10 and SARS-CoV-2 for several reasons:

- unknown factors could alter the data in unpredictable ways and only a molecular investigation will reveal the existence and implications of this phenomenon;
- the town of Codogno, where the first Lombardy outbreak occurred, recorded very high PM 10 average daily values between January 1^st^ and February 29^th^ (67 μg / m^3^) [Appendix 1];
- the novel coronavirus behavior may not be related to the daily average values of PM 10 but rather to specific thresholds, above which, its virulence and contagiousness would increase considerably. For example, an hour of very intense traffic could favor its spread much more than a constant release of the same particulate matter amount in a longer time lapse.

On the contrary, both at national level (as shown in other studies) and in Lombardy, the correlation with PM 2.5 appeared much more evident and significant. Although it is always necessary to wait for an in-depth molecular study, since there is no correlation between the number of inhabitants and PM 2.5 or between population density and PM 2.5, the correlation between PM 2.5 and SARS-CoV-2 is very likely causal in nature and far more important than that with PM 10.

## Limitations

This study is based on the results of other researches, some of which were non peer-reviewed. However, an independent verification of the data provided has been carried out. The S.E.I.R. model predictability is inversely proportional to the prediction temporal distance since the fixed values, such as basic reproduction number, incubation time, and healing time, could change in an unpredictable way; furthermore, iterative methods propagate errors divergently. Anyway, its adoption in the short term remains valid. The mortality rate is considered constant in the period February 22 - May 1, 2020. Regarding PM 10 and PM 2.5 aerosols, we only had access to cities and regions daily average values and the data of some monitoring stations were not available.

## Conclusions

The S.E.I.R. model is approximate as a direct consequence of the uncertainty of the data it was supposed to fit but it can act as a valid comparison for the identification of any SARS-CoV-2 behavior mutations, as well as for the evaluation of the containment measures effectiveness. From the comparison between the S.E.I.R predictions and the estimated real trends highlighted above, as well as between the trends of Lombardy and the other Italian regions, we report that the Italian COVID-19 data are statistically compatible with possible evolutionary mutations of SARS-CoV-2. The demographic similarity between Lombardy and Emilia Romagna, the delayed increase of COVID-19 contagiousness in Piedmont, the absence of an evident statistical correlation with daily PM 10 concentrations and nothing more than a moderate correlation with daily PM 2.5 concentrations, are sufficient reasons to assert the locality of SARS-CoV-2 behavior is also due to factors that are currently unknown. Further investigation is needed.

## Data Availability

The authors confirm that the data supporting the findings of this study are available within the article [and/or] its supplementary materials.

## Notes

### Competing Interest Statement

The authors have declared no competing interest.

### Funding Statement

No external funding was received.

